# A longitudinal study of immune cells in severe COVID-19 patients

**DOI:** 10.1101/2020.06.16.20130914

**Authors:** Didier Payen, Maxime Cravat, Hadil Maadadi, Carole Didelot, Lydia Prosic, Claire Dupuis, Marie-Reine Losser, Marcelo De Carvalho Bittencourt

## Abstract

Little is known about the time-dependent immune responses in severe COVID-19. Data of 15 consecutive patients were sequentially recorded from intensive care unit admission. Lymphocyte subsets and total monocyte and subsets counts were monitored as well as the expression of HLA-DR. For 5 patients, SARS-CoV-2-specific T-cell polyfunctionality was assessed against Spike and Nucleoprotein SARS-CoV-2 peptides. Non-specific inflammation markers were increased in all patients. Median monocyte HLA-DR expression was below the 8,000 AB/C threshold defining acquired immunodepression. A “V” trend curve for lymphopenia, monocyte numbers, and HLA-DR expression was observed with a nadir between days 11-14 after symptoms’ onset. Intermediate CD14^++^CD16^+^ monocytes increased early with a reduction in classic CD14^++^CD16^-^ monocytes. Polyfunctional SARS-Cov-2-specific CD4 T-cells were present and functional, whereas virus-specific CD8 T-cells were less frequent and not efficient. We report a temporal variation of both innate and adaptive immunity in severe COVID-19 patients, helpful in guiding therapeutic decisions (e.g. anti-inflammatory *vs*. immunostimulatory ones). We describe a defect in virus-specific CD8 T-cells, a potential biomarker of clinical severity. These combined data also provide helpful knowledge for vaccine design.

**Trial registration number:** NCT04386395

## INTRODUCTION

The SARS-CoV-2 outbreak causes a spectrum of clinical patterns that vary from asymptomatic but potentially contagious infection to mildly symptomatic and severe forms (Zhou et al., 2020). The latter brings the patient to Intensive Care Units (ICU) for mechanical ventilation (Phua et al., 2020). Currently, little is known about clinical patterns and immune response relationships.

Significant modifications reported in severe forms of COVID-19 include elevated nonspecific plasma markers (C-reactive Protein [CRP], ferritin, lactate dehydrogenase [LDH]) associated with marked lymphopenia, mainly of CD4 and CD8 T-cell subsets (Chen et al., 2020b; Li et al., 2020). The latter, associated with decrease in HLA-DR expression on monocytes (Giamarellos-Bourboulis et al., 2020), suggest acquired immune-suppression, as observed in bacterial sepsis (Lukaszewicz et al., 2009). Elevated levels of pro-inflammatory cytokines, mainly IL-6, have led to the hypothesis of an innate-mediated “cytokine storm” driving neutrophilia and defective antigen-presentation (Giamarellos-Bourboulis et al., 2020). These dysregulations of both innate and adaptive immunity (Vardhana and Wolchok, 2020) explain apparent contradictions in COVID-19 therapeutic trials (Ingraham et al., 2020). A better understanding of the longitudinal evolution of innate and adaptive immune responses during severe COVID-19 disease is needed before decisions on immune-modulation and clinically relevant immune-monitoring (Payen, 2020).

This prospective study of ICU COVID patients reports a monocentric longitudinal evaluation of innate immunity based on monocyte subsets expression of HLA-DR (Vardhana and Wolchok, 2020; Wong et al., 2012) and of adaptive immunity based on lymphocyte subsets absolute numbers (AN) and functions, referring to the onset of symptoms. Notably, we report on the presence of peripheral TH1-type SARS-CoV-2-specific T-cells in ICU COVID patients.

## METHODS

### PATIENTS

From March 30 to April 30, 2020, 15 cases of confirmed COVID-19 (positive RT-PCR for SARS-CoV-2 and suggestive chest CT-Scan) were prospectively investigated after ICU admission at the University Hospital of Nancy (CHRU-Nancy), France. The protocol was approved by the Innovation and Research Direction (reference 2020PI080), and by the Research Ethical Committee (Saisine 263) of CHRU-Nancy and registered at nih.gov (NCT04386395). No additional samples were drawn and no cells or plasma were stored after completion of the study. Relatives or patients themselves were questioned about objections to use the collected data for scientific purposes and/or potential publications. These statements and non-opposition forms were dated and recorded in medical files.

After medical team consensus, patients did receive neither direct/indirect anti-viral treatment nor immunomodulating drugs except for 3 patients who received low dose steroids, allowing results interpretation based on a relatively pure COVID-19 natural evolution. Medical history, delay from the onset of symptoms and ICU admission (Table S1), classic clinical and routine biological data were recorded.

### STUDY DESIGN

Complete blood cell evaluation was categorized according to time intervals from both the onset of symptoms and ICU admission for blood sampling as follows (Figure S1): A: days (d)7-10 as a 1^st^ period (48 hours after ICU admission); B from d11-14; C from d15-18; D from d19-23; E: after d24. Similar analyses were performed referring to the delay from ICU admission: A: d0-4; B: d5-8; C: d9-12; D: late > d12 (Table S2).

### LABORATORY INVESTIGATIONS

Routine parameters, nonspecific inflammatory markers and immune-cells characterization were first measured on ICU admission (Tables 1 and 2) and these values considered as baseline. Serial measurements were repeated until the patient was discharged or died. Flow-cytometry whole-blood routine analyses of circulating monocytes and lymphocytes were performed at the Diagnostic Flow-Cytometry platform of CHRU Nancy by using the BD FACSLyric™ Clinical System (BD Biosciences, San Jose, CA). All fluorochrome-conjugated antibodies and reagents were from BD Biosciences.

**Table 1.**
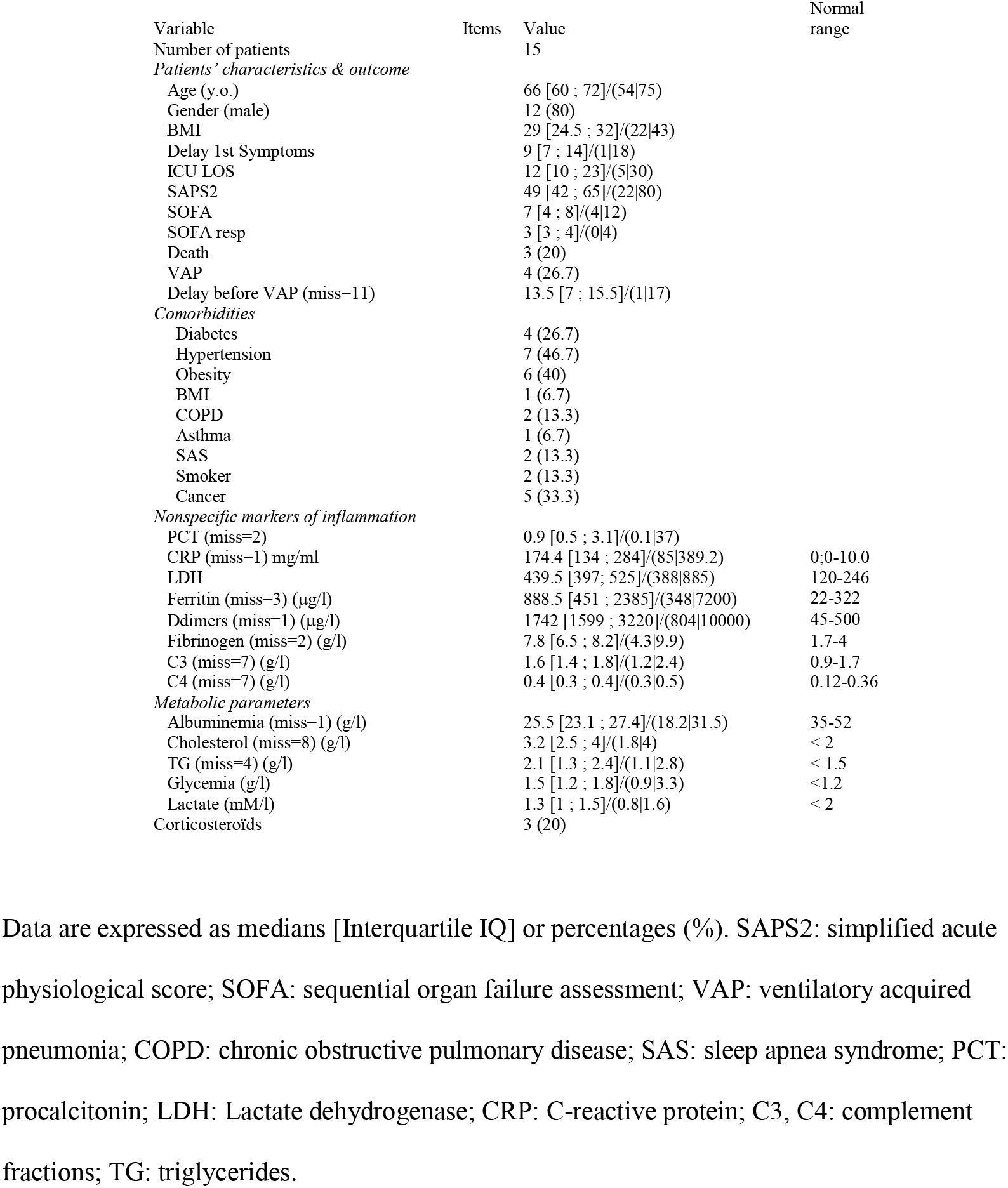
Clinical characteristics, comorbidities, severity scores, nonspecific markers, metabolic parameters at the time of enrollment.

| Variable | Items | Value | Normal range |
| --- | --- | --- | --- |
| Number of patients |  | 15 |  |
| *Patients' characteristics & outcome* |  |  |  |
| Age (y.o.) |  | 66 [60 ; 72]/(54\|75) |  |
| Gender (male) |  | 12 (80) |  |
| BMI |  | 29 [24.5 ; 32]/(22\|43) |  |
| Delay 1st Symptoms |  | 9 [7 ; 14]/(1\|18) |  |
| ICU LOS |  | 12 [10 ; 23]/(5\|30) |  |
| SAPS2 |  | 49 [42 ; 65]/(22\|80) |  |
| SOFA |  | 7 [4 ; 8]/(4\|12) |  |
| SOFA resp |  | 3 [3 ; 4]/(0\|4) |  |
| Death |  | 3 (20) |  |
| VAP |  | 4 (26.7) |  |
| Delay before VAP (miss=11) |  | 13.5 [7 ; 15.5]/(1\|17) |  |
| *Comorbidities* |  |  |  |
| Diabetes |  | 4 (26.7) |  |
| Hypertension |  | 7 (46.7) |  |
| Obesity |  | 6 (40) |  |
| BMI |  | 1 (6.7) |  |
| COPD |  | 2 (13.3) |  |
| Asthma |  | 1 (6.7) |  |
| SAS |  | 2 (13.3) |  |
| Smoker |  | 2 (13.3) |  |
| Cancer |  | 5 (33.3) |  |
| *Nonspecific markers of inflammation* |  |  |  |
| PCT (miss=2) |  | 0.9 [0.5 ; 3.1]/(0.1\|37) |  |
| CRP (miss=1) mg/ml |  | 174.4 [134 ; 284]/(85\|389.2) | 0;0-10.0 |
| LDH |  | 439.5 [397 ; 525]/(388\|885) | 120-246 |
| Ferritin (miss=3) (µg/l) |  | 888.5 [451 ; 2385]/(348\|7200) | 22-322 |
| Ddimers (miss=1) (µg/l) |  | 1742 [1599 ; 3220]/(804\|10000) | 45-500 |
| Fibrinogen (miss=2) (g/l) |  | 7.8 [6.5 ; 8.2]/(4.3\|9.9) | 1.7-4 |
| C3 (miss=7) (g/l) |  | 1.6 [1.4 ; 1.8]/(1.2\|2.4) | 0.9-1.7 |
| C4 (miss=7) (g/l) |  | 0.4 [0.3 ; 0.4]/(0.3\|0.5) | 0.12-0.36 |
| *Metabolic parameters* |  |  |  |
| Albuminemia (miss=1) (g/l) |  | 25.5 [23.1 ; 27.4]/(18.2\|31.5) | 35-52 |
| Cholesterol (miss=8) (g/l) |  | 3.2 [2.5 ; 4]/(1.8\|4) | < 2 |
| TG (miss=4) (g/l) |  | 2.1 [1.3 ; 2.4]/(1.1\|2.8) | < 1.5 |
| Glycemia (g/l) |  | 1.5 [1.2 ; 1.8]/(0.9\|3.3) | <1.2 |
| Lactate (mM/l) |  | 1.3 [1 ; 1.5]/(0.8\|1.6) | < 2 |
| Corticosteroids |  | 3 (20) |  |

Data are expressed as medians [Interquartile IQ] or percentages (%). SAPS2: simplified acute physiological score; SOFA: sequential organ failure assessment; VAP: ventilatory acquired pneumonia; COPD: chronic obstructive pulmonary disease; SAS: sleep apnea syndrome; PCT: procalcitonin; LDH: Lactate dehydrogenase; CRP: C-reactive protein; C3, C4: complement fractions; TG: triglycerides.

**Table 2.**
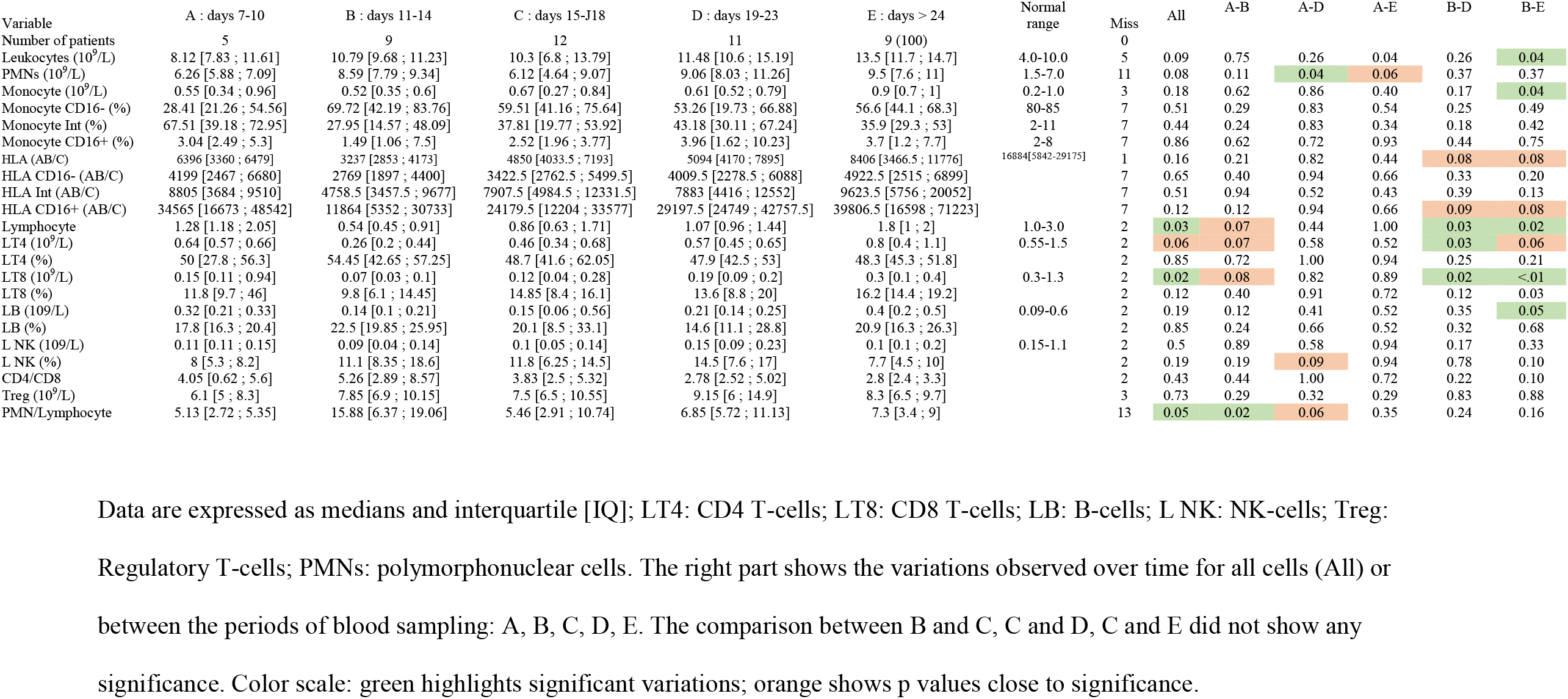
Time intervals of absolute and relative values referring to the onset of symptoms for blood lymphocytes, monocytes, polymorphonuclear cells and human leucocyte antigens-DR (HLA-DR, expressed in AB/C number of events per cell).

#### Innate immunity

Whole-blood monocytes were selected as previously reported (Figure S2) (Payen et al., 2019). CD14-positive (+) cells with intermediate SSC were analyzed. Because of their specialized functions and phenotypes (Sampath et al., 2018; Wong et al., 2012), monocyte subsets were further analyzed based on CD14 and CD16 expression as classical (CD14^++^CD16^-^), non-classical (CD14^low^CD16^++^), and intermediate (CD14^++^CD16^+^) (Ziegler-Heitbrock, 2015).

Relative and absolute populations sizes were determined and HLA-DR expression was quantified as number per cell (antibodies bound per cell [AB/C] arbitrary units) using a commercial kit (Quantibrite™, BD Biosciences). Total monocytes HLA-DR median expression in healthy donors, using similar set-up conditions, was 16,884 (5,842-29, 175) AB/C. Our laboratory’s threshold for acquired immunodepression diagnosis is 8,000 AB/C (Gouel-Cheron et al., 2015; Payen et al., 2019). “Normal proportions” for monocyte subsets are 80-95% for classical monocytes, 2-8% for non-classical monocytes and 2-11% for intermediate monocytes (Sampath et al., 2018; Ziegler-Heitbrock, 2015).

#### Adaptive immunity

Routine whole-blood immunophenotyping of T, B and NK lymphocytes by flow-cytometry determined relative and AN of lymphocytes and their subsets, i.e. CD3^+^ T-cells, CD3^+^CD4^+^ T-cells, CD3^+^CD8^+^ T-cells, CD19^+^ B-cells, CD3^-^CD16^+^56^+^ NK-cells. Regulatory T-cells (Treg) were identified in whole blood as CD3^+^CD4^+^CD25^++^CD127^neg/low^ cells (Liu et al., 2006; Seddiki et al., 2006). SARS-CoV-2-specific T-cell polyfunctionality (Boyd et al., 2015; Seder et al., 2008) against viral peptides was assessed in 5 patients to further explore COVID-19 adaptive immunity (patients #7, #9, #11, #12 and #13). Patients’ PBMC from 19 to 29 days after the onset of symptoms were stimulated *ex-vivo* by overlapping peptides covering protein sequences of the Spike and Nucleoprotein SARS-CoV-2 antigens (JPT Peptide Technologies, Berlin, Germany). Antigen-specific T-cells reactivity was assessed by intracellular IFN-γ, TNF-α and IL-2 production (Lamoreaux et al., 2012). Navios^®^ flow-cytometer and Kaluza^®^ software v2.1 (Beckman Coulter, Miami, FL) were used for data acquisition and analysis. As positive controls, PBMC were also stimulated by a mix of immunodominant microbial peptides (CEFX peptide pool; JPT Peptide Technologies) for diverse microbial antigens-specific TH1 responses. Antigen-specific CD4 and CD8 T-cells polyfunctionality was expressed by using the “Polyfunctionality Index” (PI) (Boyd et al., 2015; Larsen et al., 2012a). More details in reagents, flow-cytometry gating strategy used for the selection and quantification of Sars-Cov-2-specific-T-cells are shown in Table S3 and Figure S3.

### STATISTICAL ANALYSES

Data are described as number (%) and median (interquartile range (IQR)) for categorical and continuous variables respectively. Comparisons relied on Fisher exact test for categorical data; Kruskal-Wallis or Wilcoxon tests were used for continuous data and Wilcoxon test for paired data test whenever necessary. An ANCOVA test for repeated measurements was used for comparison of the different types of HLA-DR. A p-value of less than 0.05 was considered statistically significant. All analyses were performed using SAS software, version 9.4 (SAS Institute Inc., Cary, NC).

## RESULTS

### COHORT OF CONSECUTIVE PATIENTS

Fifteen severe COVID-19 ICU patients, median age 66 years old [60-72], 80% male, were investigated (Table 1). Because of hypoxia deterioration, all except 1 (Table S1) needed intubation and mechanical ventilation with a protective lung protocol. Importantly, none of the patients received any direct or indirect antiviral drugs nor specific immunomodulating drugs. Only 3 patients received low dose of prednisolone during their ICU stay (Table 1). Comorbidities were present in 47% of the patients (Table 1). Hypertension (46.7%) and obesity (40%) were the most frequent, followed by pre-existent cancer (33.3%) and diabetes (26.7%) (Table 1). Clinical severity was high with a median Simplified Acute Physiology Score (SAPS)2 at 49 [42-65] and a global Sepsis-related Organ Failure Assessment (SOFA) at 7 [4-8] (Table1). Three patients died (20%) during the 2^nd^ or 3^rd^ week after ICU admission. All other patients were ICU discharged. A ventilator-acquired pneumonia according to classical CDC definition (Calandra et al., 2005) was diagnosed in 26.7% of the patients.

### LABORATORY FINDINGS

Non-specific markers of systemic inflammation such as CRP, ferritin, DDimers, fibrinogen, LDH, and the complement fraction C4 were largely above normal ranges in 100% of the patients at ICU admission (Table 1). DDimers levels were 3 times above the highest normal limit.

Glycemia was slightly superior to normal values (1.5 [1.2; 1.8] g/L), with normal lactate levels (1.3 [1 ; 1.5]mM/L). Surprisingly, cholesterol (3.2 [2.5 ; 4] g/L) and triglycerides (2.1 [1.3 ; 2.4] g/L) levels were also largely above normal ranges, a lipid abnormality never reported in severe septic patients (Biller et al., 2014). AN and relative values of leukocyte subsets are shown in Table 2 and Figure 1. Considering the delay stratification from symptoms onset, there was globally no significant change in leukocyte AN (p= 0.09). Polymorphonuclears (PMN) initial AN were lower than the median observed at periods “D” (p = 0.04) and “E” (p = 0.06) (Table 2 and Figure 1). A difference in monocyte AN was only significant when phase B (d11-14) was compared to the late phase E (p = 0.04) (Table 2 and Figure 1). Importantly, the relative AN for the 3 major subtypes of monocytes did not change over time: CD16^+^-intermediate cells proportions at ICU admission were largely above the admitted proportions (Ozanska et al., 2020; Wong et al., 2012), with a concomitant reduction in the CD16^-^ cell subset. The ability of monocytes to express HLA-DR was lower than the threshold for acquired immunosuppression diagnosis at all periods, except the latest one (>24 days after initial symptoms) (Figure 2). The lowest level of immunosuppression was observed at phase B and tended to recover during the ICU stay (p = 0.08) (Figure 2). Similar patterns were observed for all monocyte subtypes, albeit more pronounced for CD16^+^ ones (non-classical and intermediate). A significant positive correlation was found at all periods between HLA-DR expression and T-cells AN for CD4 (R^2^ = 0.28; p <0.01) and CD8 T-cells (R^2^ =0.22; p < 0.01) (not shown). Also, significant linear positive correlations were seen between HLA-DR expression on intermediate and non-classic monocyte subsets and CD4 T-cells AN (p <0.01) and between CD16^+^ intermediate subset and CD8 T-cells AN (p<0.01) (not shown). Considering alive of dead patients as those above or below the median SAPS2, no differences were observed for monocyte subsets proportions.

**Figure 1:**
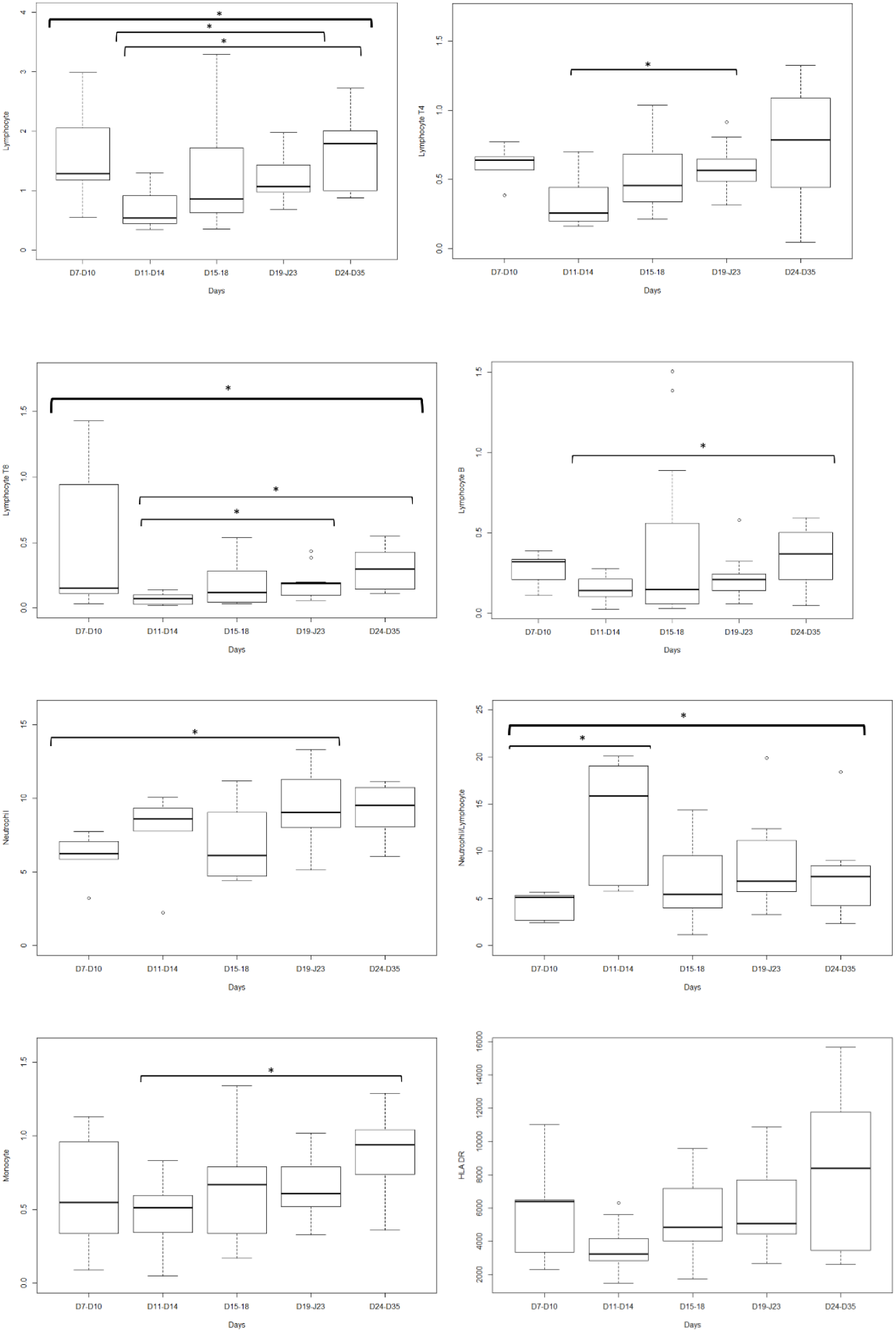
Leukocyte absolute number variations according to the delay from the onset of symptoms. HLA-DR: Human Leucocyte Antigen-DR expressed as events per cell (AB/C); * p<0.05

**Figure 2:**
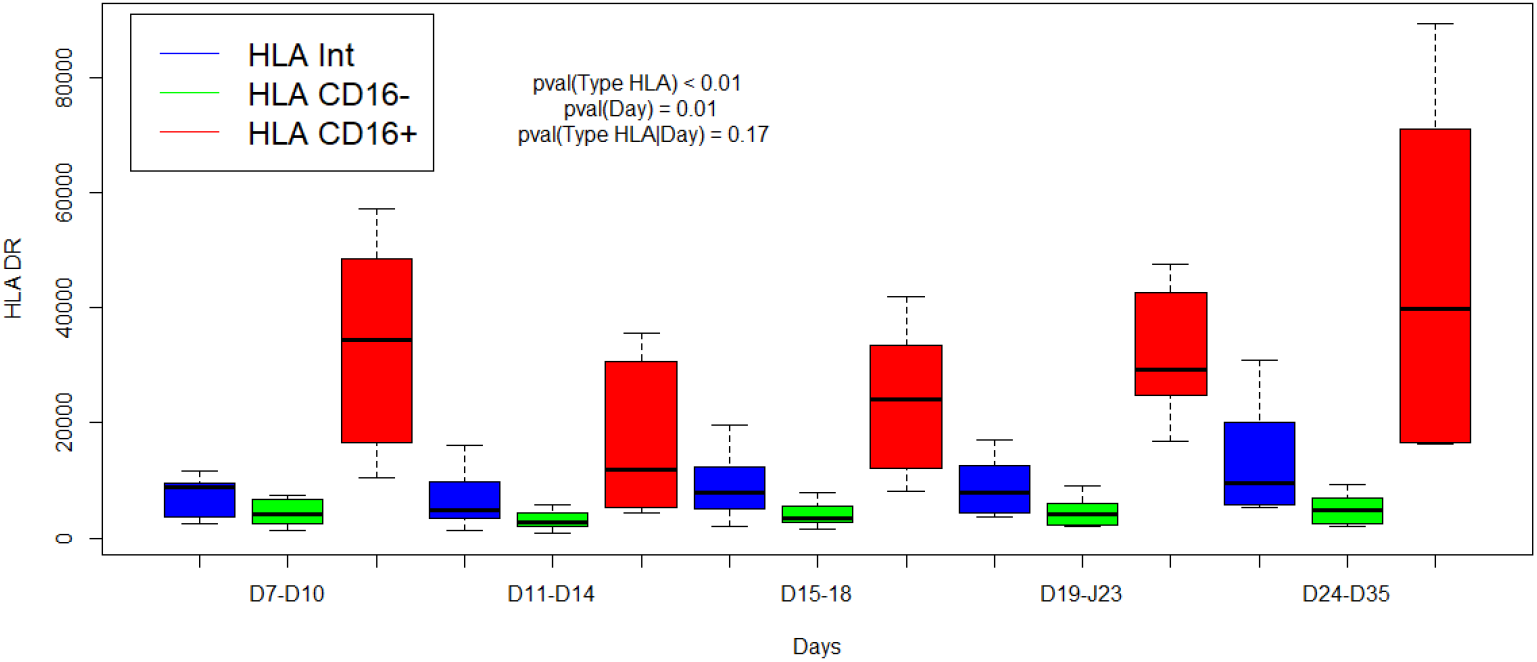
Time evolution of HLA-DR expression according to the delay from the onset of symptoms for 3 monocyte subtypes. Green: classical (CD14^++^CD16^-^) monocytes; red: non-classical (CD14^+^CD16^++^), monocytes; blue: intermediate (CD14^++^CD16^+^) monocytes. HLA-DR: Human Leucocyte Antigen-DR expressed as events per cell (AB/C).

As reported (Chen et al., 2020a), global lymphocyte counts showed a severe reduction in AN compared to normal ranges (Table 2 and Figure 1). Lymphocyte AN changed significantly over time (p = 0.03). CD4 (“B” vs “D”: p = 0.03) and CD8 (“B” vs “D”: p = 0.02) T-cell AN were reduced at period B, being the “nadir” of lymphopenia, and then followed by a slow CD4 and CD8 T-cells increase (p<0.03) (Figure 1). The CD4/CD8 ratio was not impacted by lymphopenia. B-cells, NK-cells, and Treg numbers were stable along the monitoring time (Table 2). The leukocyte moderate elevation associated with the total lymphocytes rapid decline resulted in a significant rise in PMNs/Lymphocytes ratio at the “nadir” period B, with subsequent normalization (Table 2).

### FUNCTIONAL ANALYSIS OF CELLULAR IMMUNE RESPONSE IN SARS-CoV-2 PATIENTS

Peripheral SARS-CoV-2-specific T-cells were identified in 5 patients by using an intracellular staining assay and flow-cytometry to evaluate the production of 3 TH1 cytokines contributive to viral clearance (Seder et al., 2008). PBMC were stimulated by 3 SARS-CoV-2 peptide pools (Sk1 and Sk2 pools and Nucleoprotein (NC pool)). The polyfunctionality Index (PI) for IFN-γ, TNF-α and IL-2 production showed that virus-specific CD4 T-cells were more polyfunctional than CD8 T-cells (Figure 3). Remarkably, a high proportion of bi-functional and tri-functional CD4 T-cells was observed, whereas CD8 T-cells were essentially monofunctional (Figure 3). Nucleoprotein-specific CD4 T-cells were less numerous than those specific for the Spike glycoprotein (both Sk1 and Sk2 pools), similarly to reported results (Weiskopf et al., 2020). CD4 T-cells responses to immunodominant peptides from different infectious agents (CEFX pool) were much lower than CD8 T-cells responses (Figure 4).

**Figure 3:**
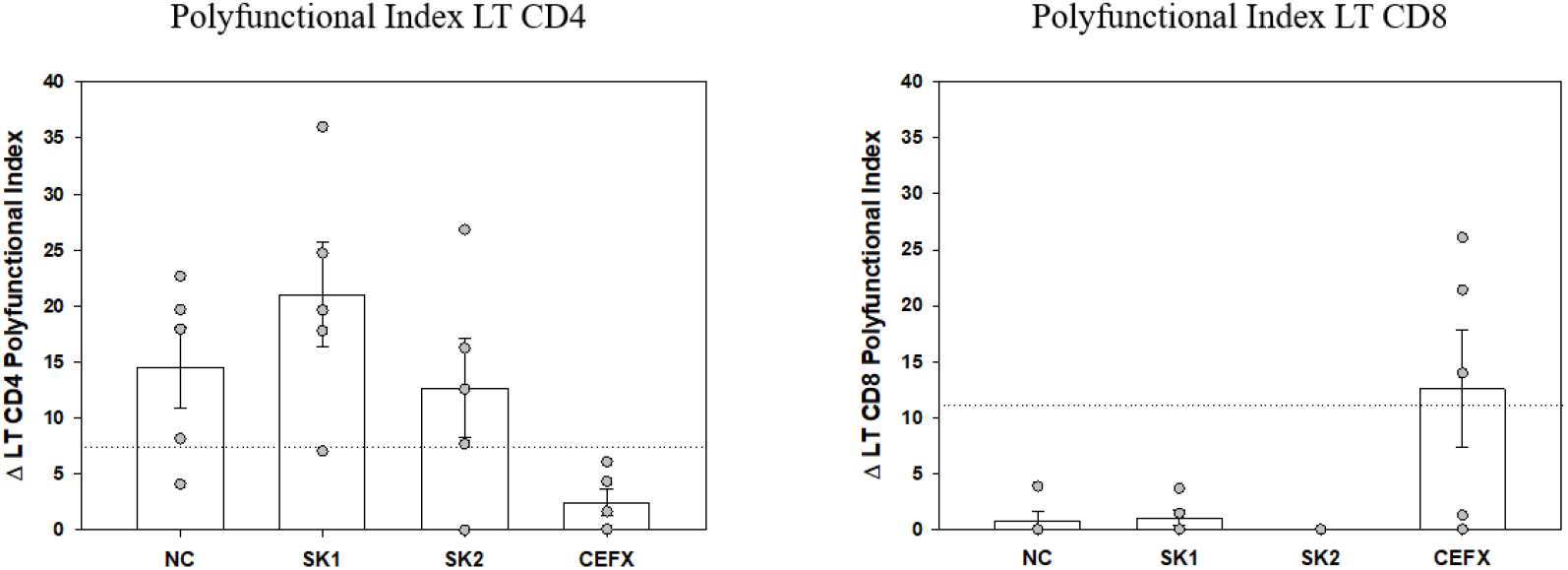
Polyfunctional Index (PI) of SARS-CoV-2-specific CD4 and CD8 T-cells. The PI of antigen-specific CD4 and CD8 T-cells from 5 patients was calculated as previously described (Boyd et al., 2015; Larsen et al., 2012a) for each antigen stimulation. Data are expressed after subtraction of “background polyfunctionality” of medium-stimulated cells (Δ PI). Dotted lines correspond to PI positivity threshold as determined at 3SD of 30 negative controls measures for CD4 and CD8 T-cells. Bars are shown as mean ± SEM. LT CD4: CD4 T-cells, LT CD8: CD8 T-cells.

**Figure 4:**
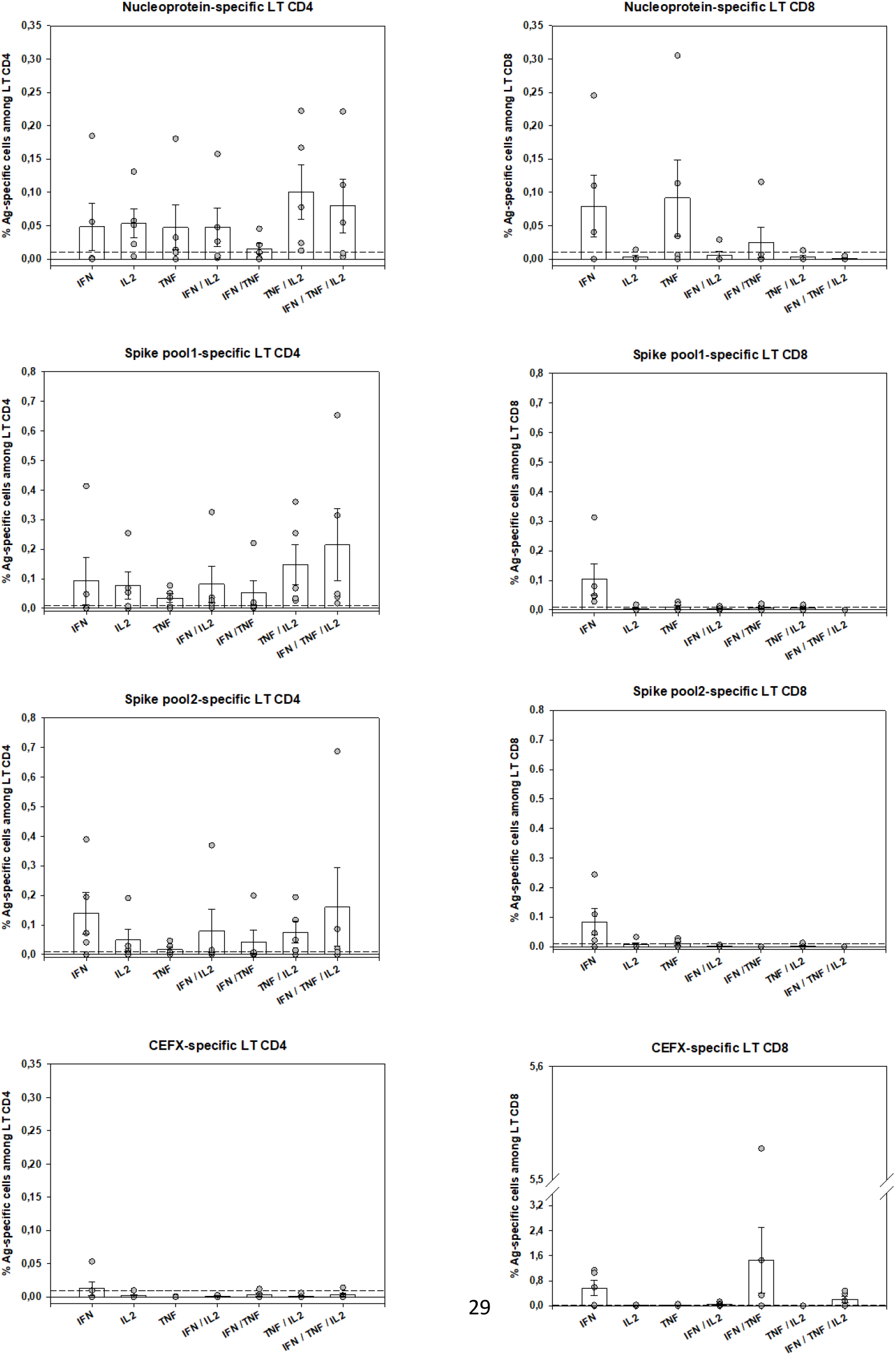
SARS-CoV-2-specific-T-cells numbers and functions. Peripheral antigen-specific CD4 and CD8 T-cells from 5 patients were identified by intracellular staining of IFN-γ, TNF-α and IL-2 by flow-cytometry as described (see Material and Methods and Suppl. Methods sections). Shown are individual data from stimulation with SARS-CoV-2 Nucleoprotein and Spike Glycoprotein peptides pools as well as by multiple immunodominant microbial peptides (CEFX pool) for CD4 and CD8 T-cells and for each cytokine capability (monofunctional, bifunctional or trifunctional). Basal cytokine production of medium-stimulated cells was subtracted for each antigen-stimulated cell subtype and a 0.01% positivity threshold was defined (dotted lines). Bars are shown as mean ± SEM. LT CD4: CD4 T-cells, LT CD8: CD8 T-cells.

## DISCUSSION

The most significant predictors of disease severity in COVID-19 infection relate to both innate and adaptive immunity and their inter-relations. Until now, to our knowledge, no longitudinal clinical study on ICU-admitted patients has reported the evolution of monocyte subsets with concomitant HLA-DR expression and lymphocytes subsets AN. Precise knowledge of symptoms’ onset allowed us to interpret the collected data as related to disease progression. Between symptoms’ onset and ICU admission to ICU discharge or death, we observed a “V” curve trend for monocytes and their HLA-DR expression as well as for lymphopenia, with a nadir between days 11 to 14. This 2 weeks timing already reported as a risk period for clinical events (Zhou et al., 2020) was characterized by the lowest HLA-DR expression on monocyte subsets associated with the deepest CD4 and CD8 T-cells lymphopenia. We found that mainly polyfunctional SARS-Cov-2-specific CD4 T-cells were present 3 weeks after the onset of symptoms, while peripheral SARS-Cov-2-specific CD8 T-cells were less numerous and not efficient. However, a normal CD8 T-cells response for other microbial antigens was present, excluding a global functional defect of peripheral CD8 T-cells.

This ICU cohort had a predominance of males with high Body Mass Index and frequent comorbidities, as previously reported (Guan et al., 2020). Acute respiratory syndrome with severe hypoxia imposed intubation and mechanical ventilation for 14/15 patients. However, our cohort differs from others by several aspects: no patient received any anti-viral therapy before and during ICU stay, avoiding a potential incidence on the immune profile. Moreover, no patient received specific immunomodulation therapy. At referral to ICU, all patients had elevated levels of non-specific markers of systemic inflammation such as ferritin, LDH, and CRP. The mortality rate (20%) was relatively low and none of the patients experienced shock nor multiorgan failure. The reasons for that are not clear, except the reduced interference between drugs and immune status.

In absence of a longitudinal assessment, one point checking for immune status has big limitations. As shown here, both innate and adaptive immunity vary over time after SARS-CoV-2 infection. COVID-19 severity is related to an initial excessive inflammatory response, pro-inflammatory cytokine storm and global lymphopenia as well as pulmonary mononuclear cell infiltration (Xu et al., 2020). The reported monocyte and macrophage hyperactivation have a major role on this hyperinflammatory state, potentially depending on their interaction with virus-specific T-cells (Giamarellos-Bourboulis et al., 2020; Merad and Martin, 2020; Tay et al., 2020). The virus itself, directly via pathogen-associated molecular patterns and indirectly via damage-associated molecular patterns, may activate multiple immune pathways (Vardhana and Wolchok, 2020).

If monocytes can initiate and amplify adaptative immune responses, they also play a key role supporting tissue homeostasis by resolving these responses to avoid excessive tissue damage (Wong et al., 2012). Monocyte classical (CD14^++^CD16^-^), non-classical (CD14^+^CD16^++^) and intermediate (CD14^++^CD16^+^) subsets reflect different functions (Ozanska et al., 2020; Wong et al., 2012). Only one study has reported monocyte subsets proportions in 3 ICU COVID-19 patients (Zhang et al., 2020). In our study, monocyte AN did not change along with COVID evolution in ICU, except when comparing the last measurements (>24 days) with those of days 11-14 after symptoms onset. Previous reports also showed maintained monocyte numbers (Merad and Martin, 2020). Proportions of the 3 monocyte subsets did not change significantly during the evolution period we studied. However, the intermediate CD14^++^CD16^+^ subset was remarkably always above reported normal values (Ozanska et al., 2020; Wong et al., 2012), which was associated with reduced proportions of classic CD14^++^CD16^-^ monocytes, as reported (Zhang et al., 2020). Functionally, HLA-DR global expression was below the 8,000 AB/C threshold defining acquired immunosuppression (Gouel-Cheron et al., 2015; Payen et al., 2019), with similar trends for the 3 subsets. Such changes in monocyte proportions and functionality appear related to disease severity and suggest a maturation towards macrophages (Merad and Martin, 2020). The severe reduction in HLA-DR expression observed was comparable with that observed in bacterial sepsis (Lukaszewicz et al., 2009) or trauma (Gouel-Cheron et al., 2015). The reported SARS-CoV-2 virus-induced restriction in interferon genes expression may account for severe reduction in different IFN proteins, including IFN-γ (Tay et al., 2020). The close correlation between CD4 and CD8 T-cells numbers and monocyte HLA-DR expression levels supports such hypothesis. The severe reduction in monocyte HLA-DR expression may also result from other mechanisms such as monocyte dysfunction, particularly secondary to exposure to IL-6 (Giamarellos-Bourboulis et al., 2020; Wang et al., 2020), which requires further investigation.

COVID-19 mortality has been shown to correlate to global T-cell function as indirectly measured by T-cell lymphopenia (Zeng et al., 2020). However, the successful containment of viral infections depends on the generation of antigen-specific TH1-polarized CD4 and CD8 T-cells, with a huge increase of effector T-cells at acute phase, which declines after successful virus control, yet maintaining in increased pool of pathogen-specific memory T-cells (Cox et al., 2013; Swain et al., 2012). More than the quantity itself, anti-viral “T-cells quality” is widely recognized as the most important for successful anti-viral responses. It relates to the cells’ capacity to perform simultaneous anti-viral functions (e.g. cytokine or chemokine production, cytotoxicity, proliferation) (Boyd et al., 2015; Seder et al., 2008). Indeed, polyfunctional and monofunctional T-cells differ at molecular levels (Burel et al., 2017). At the time of writing, only 4 publications have assessed SARS-CoV-2-specific T-cells. Ni *et al* described them in COVID-19 recovered patients, using only IFN-γ ELISpot, possibly underestimating the magnitude and breadth of the response and not discriminating CD4 or CD8 virus-specific T-cells (Ni et al., 2020). The other 3 studies identified by flow-cytometry T-cells upregulating activation markers (Braun et al., 2020; Grifoni et al., 2020; Weiskopf et al., 2020). Although this “global activation” approach can potentially detect all antigen-specific T-cells, polyfunctional CD4 T-cells were not investigated. Grifoni *et al*. described only polyfunctional CD8 T-cells, assessed from recovered patients (Grifoni et al., 2020). Therefore, to date no study has assessed both CD4 and CD8 T-cell polyfunctionality in the COVID-19 context, especially on ICU patients. We believe that such evaluation is important to better understand immune responses against this new virus. Of note, in previous SARS-CoV epidemics, specific polyfunctional CD4 and CD8 T-cells were present several years after infection (Oh et al., 2011). The large combinatorial datasets of multiparametric flow-cytometry analysis of polyfunctionality can be integrated in a one-dimensional numerical tool, called polyfunctional index (PI) that integrates degrees and variations of cellular polyfunctionality (Boyd et al., 2015; Larsen et al., 2012a). Its application here showed that polyfunctional CD4 T-cells were present for at least one of the SARS-CoV-2 tested antigens in all 5 tested patients. Conversely, CD8 polyfunctional T-cells were not identified for any SARS-CoV-2 antigen in any patient. SARS-CoV-specific CD4 T-cells were mainly bi-(mostly TNF-α+IL-2+) and tri-functional (plus IFN-γ). Conversely, when present, the rare SARS-CoV-2-specific CD8 -reactive T-cells were mainly monofunctional (producing either IFN-γ or TNF-α). Of note, the CD8 T-cells reactive to the control mix of microbial peptides mentioned above (CEFX pool) were perfectly polyfunctional, indicating again that only the pool of peripheral SARS-CoV-2-specific CD8 T-cells is affected (Figures 3 and 4). This difference in quantity and quality between CD4 and CD8 SARS-CoV-2-specific T-cells might be explained by a preferential homing of the virus-specific effector CD8 T-cells to tissues, and especially the lungs and/or by T-cell functional exhaustion (Diao et al., 2020; Wherry and Kurachi, 2015). The latter hypothesis, that would imply selective PD-1 expression, is not supported by the good specific responses of helper CD4 T-cells in the same samples. Further evaluations of SARS-CoV-2-specific T-cells in bronchoalveolar lavage and of the expression of T-cell exhaustion inhibitory markers (e.g. PD-1 and TIM-3) are then warranted. Virus-specific CD8 T-cells are key in eliminating virus-infected cells (Cox et al., 2013) and Grifoni *et al*. have reported their presence in recovered patients (Grifoni et al., 2020). Combined with our findings, one may therefore hypothesize that inefficient SARS-CoV-2-specific CD8 T-cell responses could promote viral persistence and then virus-induced inflammatory damage in severe COVID-19 patients. Overall, SARS-CoV-2-specific polyfunctional CD4 T-cells were present at relatively high numbers, by comparison to other antigen-specific T-cells routinely investigated in our laboratory (Dekeyser et al., 2017), suggesting a strong immune response. Further studies are needed to establish the kinetics of circulating SARS-CoV-2-specific T-cells over time and their relationship with COVID-19 clinical presentation.

In conclusion, we report the first concomitant and longitudinal evaluation of innate and adaptive immunity in COVID-19 severe cases. We believe that such extensive immunomonitoring studies are needed in order to accumulate a better knowledge in SARS-CoV-2 innate and adaptive immune-responses relationships. This approach might be helpful in making therapeutic decisions (e.g. anti-inflammatory *vs*. immunostimulation interventions) depending on the stage of the disease. This also will be of utmost importance for vaccine efficacy evaluation in future clinical trials.

## Data Availability

Availability of data referred to in the manuscript will be evaluated on an each individual basis.

## Acknowledgements

The authors thank the helpful comments of Professors Philippe Saas (Besançon, France) and Marie-Christine Béné (Nantes, France) as well as the support of Drs. Patricia Franck and Véronique Latger-Cannard from Pôle Laboratoires of CHRU-Nancy. The authors also thank the paramedical and medical team taking care of COVID-19 patients in the Surgical Intensive Care Unit of CHRU-Nancy at Brabois Hospital.

## Supplement

### Supplementary methods

#### Flow-cytometry assessment of SARS-CoV-2 specific T-cells

Peripheral blood was obtained by venipuncture and collected in heparin-containing tubes (BD Biosciences). Mononuclear cells (PBMC) were separated by density gradient no later than 6 hours after venipuncture, washed and counted prior to cell culture. PBMC (5⨯10^5^ to 1.5⨯10^6^ cells) were resuspended in 200µL of serum-free AIM-V medium and were separately stimulated overnight (12-14 hours) in 96 wells plate with peptide pools of overlapping 15-mer peptides (11 amino acid overlaps) spanning the entire Spike (divided in two pools of peptides: “SK1 and SK2”) and Nucleoprotein (pool “NC”) SARS-CoV-2 proteins (1.25 µg/mL of each peptide). Brefeldin A (Sigma-Aldrich) was added to each well and the plates set at 37°C, 5% CO_2_ humidified atmosphere. Unstimulated PBMCs (medium +DMSO, vehicle of the peptide pools) were used as negative controls. For positive controls of detecting antigen-specific responses, PBMC were separately stimulated with a control pool of 176 known immunodominant peptide epitopes of diverse infectious agents (CEFX pool). All peptide pools were from JPT Peptides (Berlin, Germany). All samples were also co-stimulated with antibodies directed against CD28 and CD49d (1 µg/mL, BD Biosciences). As positive controls of global TH1-immunocompetence, PBMC were also stimulated in parallel with Phorbol-Myristate-Acetate (50 ng/mL, Sigma-Aldrich) and Ionomycin (500 ng/mL, Sigma-Aldrich). After stimulation, cells were collected and washed before an incubation with BD Horizon™ Fixable Viability Stain 510 (FVS 510) and a cell surface antibody mix of anti-CD3 BB515, anti-CD4 PE-Cy7 and anti-CD8 PE-CF594. Cells were then fixed, washed and permeabilized according to the manufacturer’s instructions by using IntraStain® kit (Dako). Next, the cells were incubated with an intracellular antibody mix of anti-IL2 PE, anti-TNF-α APC and anti-IFN-γ BV421. All conjugated antibodies were from BD Biosciences. After 2 washes, cells were resuspended in PBS and immediately proceeded to flow cytometry data acquisition. Data were acquired using a Navios® Flow Cytometer (Beckman Coulter). SARS-CoV-2 -specific T-cells were identified using Kaluza® software v2.1 (Beckman Coulter) as by a specific gating strategy (Suppl. Figure3). The same gating strategy was used for SARS-CoV-2 Ag-stimulated cells and the percentage of Ag-specific CD4 or CD8 T cells was determined after subtracting the percentage of cytokine-positive events in unstimulated controls. Responses above the level of difference of 0.01% cytokine-producing cells were considered as Ag-specific positives (PMID: 19266489). In order to quantify the polyfunctionality of SARS-CoV-2 specific T-cells, we calculated the Special Polyfunctionality Index (PI) for each stimulation condition and for CD4 and CD8 T cells as described (Larsen et al., 2012b). We have used a higher ponderation for increasing polyfonctionnality (q=1.2) as determined in a series of statistical analysis correlating T-cell polyfunctionality and efficacy (Boyd et al., 2015). Negative controls stimulation (medium plus co-stimulatory antibodies) present a background production of cytokines (mostly monofunctional cells), we then subtracted the Polyfunctional Index of negative controls from each specific antigenic stimulation in order to correct for this “background polyfunctionality” for each patient tested. The limit of detection for an augmentation of polyfunctionality was determined at 3 SD of results from 30 measures of negative controls for CD4 and CD8 of different subjects.

**Figure S1.**
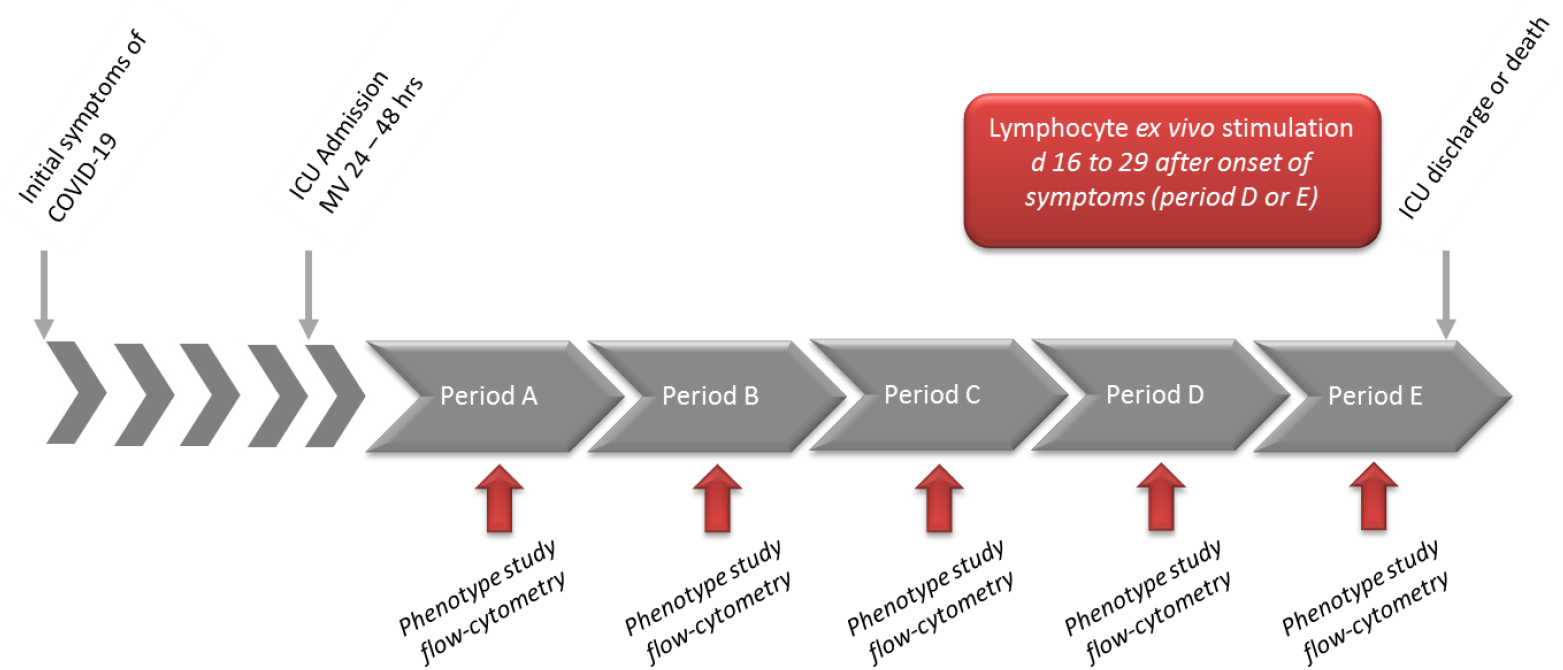
Study design of sequential monocytes and lymphocytes characterization using flow-cytometry techniques. Period A (d7-10), Period B (d11-14), Period C (d15-18), D (d19-23), Period E: > d24 after onset of disease. Ex vivo stimulation of lymphocytes (PBMC) with SARS-CoV-2 antigens was performed between d16 to d29. hrs: hours.

**Figure S2.**
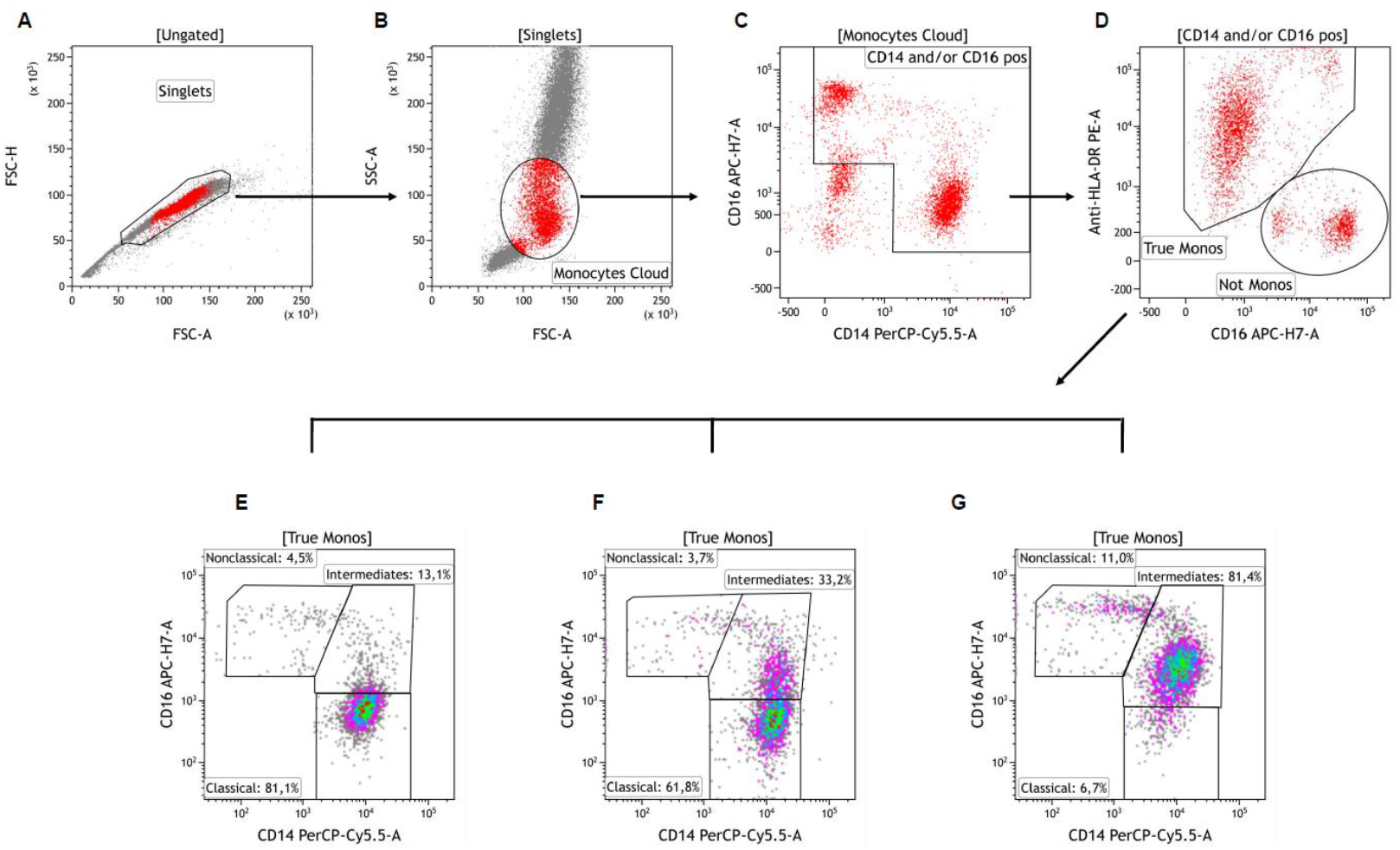
Flow-cytometry sequential gating strategy for identification of circulating monocytes subsets. Singlets were first identified on a FSC-A vs. FSC-H dot plot (Plot A). Cells were next visualized on FSC vs. SSC plot and an ample “Monocytes Cloud” gate was set, excluding most neutrophils and lymphocytes (Plot B). These cells were then viewed on a CD14 vs. CD16 plot and a gate was drawn excluding CD14 negative and CD16 low/negative cells (gate “CD14 and/or CD16 pos”, Plot C). Selected cells were next viewed on a CD16 vs. HLA-DR plot and the contaminating cells (‘‘Not Monos’’ gate) were easily distinguished from the ‘‘true’’ monocytes (“True Monos” gate, Plot D). These latter ‘‘cleaned’’ monocyte population was next viewed again on a CD14 vs. CD16 plot and gated as CD14++CD16-“Classical”, CD14++CD16+ “Intermediates” and CD14+CD16++ “Nonclassical” monocytes subsets. Plots E, F and G show representative examples of patients with low (Plot E), moderate (Plot F) and high (Plot G) proportions of Intermediate monocytes. Adapted from Abeles *et al*. (Abeles et al., 2012).

**Figure S3.**
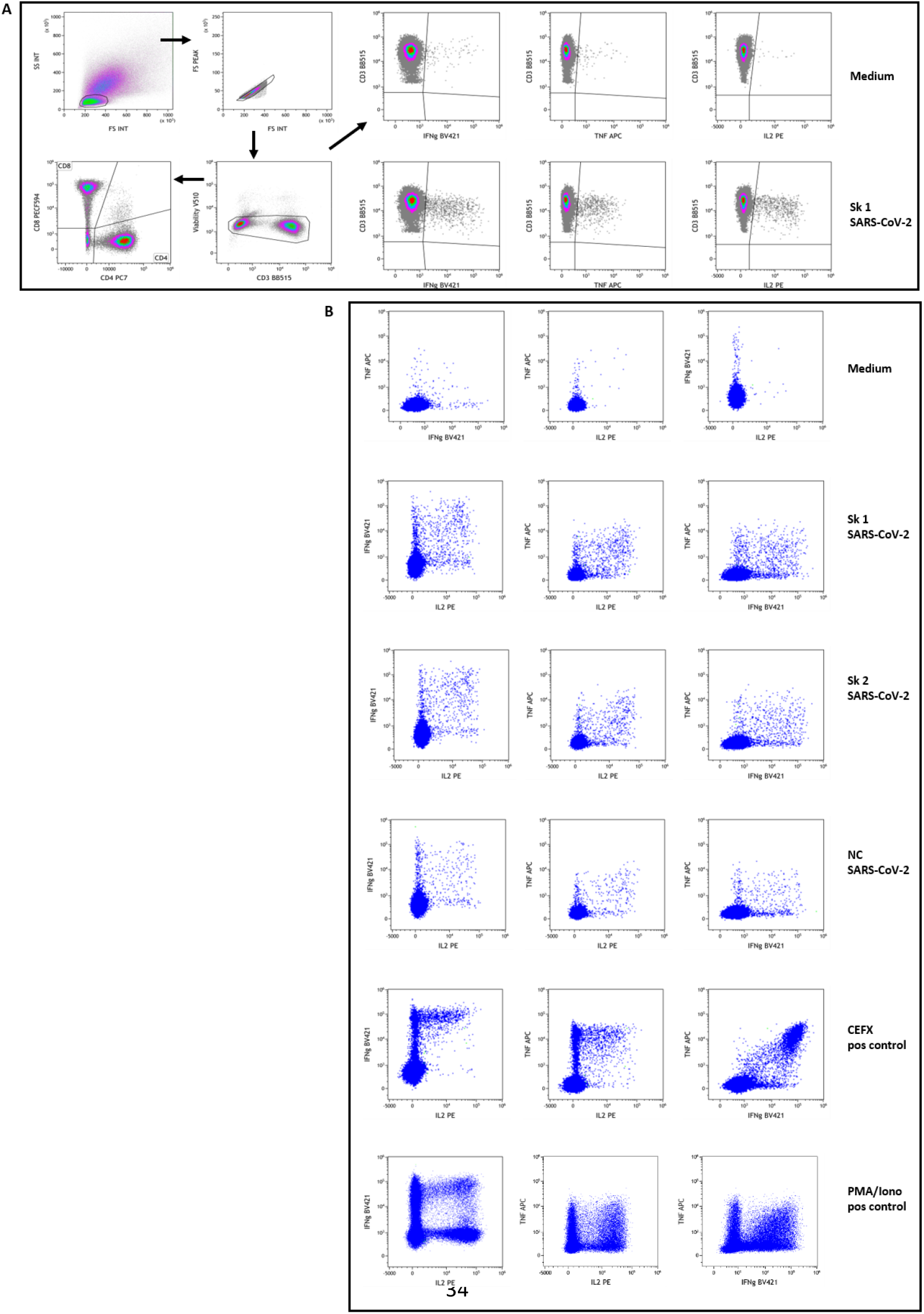
Flow-cytometry assessment of SARS-CoV-2 specific T-cells. Patients’ PBMC were stimulated overnight and then stained with fluorescent-conjugated antibodies to surface and intracellular molecules as described in Material and Methods section. Figure shows the gating strategy used to identify SARS-CoV-2 specific CD4 and CD8 T-cells in a representative subject (patient #13). Panel A: Live lymphocytes were defined after a sequential gating on FSC vs SSC, singlets and low FVS 510 staining. A gate was then set using the negative controls (medium stimulated cells) for the baseline expression of each cytokine as well as CD4 and CD8 subpopulations on total live CD3+ T lymphocytes. For cytokine-secretion assessment, boolean gates (not shown) were used to determine the frequencies of single-, double-, and triple-cytokine secreting CD4 and CD8 T-cells, defined as a percentage of IL2+, TNFα+ or IFNγ+ cells among selected T lymphocytes. These gates and analysis strategy were then set for all stimulation conditions for each patient assessed. Examples of stimulation with medium and peptides pool 1 of Spike SARSCoV-2 (Sk1) are shown at upper right. Panel B: Example dot-plots of bifunctional total T-cells identified are shown for medium (negative control), SARS-CoV-2 peptides pool 1 and 2 of Spike (Sk1 and Sk2) and Nucleoprotein (NC), as well as CEFX (multiple immunodominant microbial peptides pool) and PMA/Ionomycin positive controls stimulation. All shown data are from patient #13.

**Table S1.** Context of COVID-19 disease of the patients enrolled in the study. Table S1: Note the precise delay between the onset of symptoms and the ICU admission and intubation. ICU: intensive care unit; LOS: length of stay; MV: mechanical ventilation.

| *Patient*<br>*N°* | *Age*<br>*(range*<br>*Years)* | *Delay first symptoms -*<br>*intubation (days)* | *Overall*<br>*duration of MV*<br>*(days)* | *LOS ICU*<br>*(days)* | *Pulmonary embolism* | *Outcome*<br>*Alive = A. Dead = D* |
| --- | --- | --- | --- | --- | --- | --- |
| #1 | 50-55 | 8 | 8 | 10 | no | A |
| #2 | 60-65 | 7 | 12 | 18 | no | A |
| #3 | 60-65 | 9 | 3 | 10 | no | A |
| #4 | 70-75 | 9 | 10 | 12 | no | A |
| #5 | 65-70 | 8 | 27 | 30 | no | A |
| #6 | 65-70 | 5 | 9 | 11 | no | A |
| #7 | 70-75 | 8 | 17 | 25 | no | A |
| #8 | 60-65 | 16 | 18 | 21 | yes | A |
| #9 | 60-65 | 1 | 23 | 23 | yes | D |
| #10 | 70-75 | 14 | 14 | 14 | no | D |
| #11 | 60-65 | 11 | 6 | 9 | no | A |
| #12 | 55-60 | 5 | 21 | 24 | yes | A |
| #13 | 60-65 | 16 | 22 | 11 | no | D |
| #14 | 70-75 | 11 | 0 | 5 | no | A |
| #15 | 70-75 | 18 | 4 | 5 | no | A |

**Table S2.** Longitudinal immune parameters referring to the day of admission in Intensive Care Unit.

| Variable | A: D 0 -4 | B: D 5-8 | C: D 9-12 | D : > D12 | Normal range | All | A-B | A-C | A-D | B-C | B-D | C-D |
| --- | --- | --- | --- | --- | --- | --- | --- | --- | --- | --- | --- | --- |
| Number of patients | 9 | 12 | 12 | 6 |  |  |  |  |  |  |  |  |
| Leucocytes | 1.02 [0.53 ; 1.28] | 0.84 [0.55 ; 1.18] | 1.06 [0.91 ; 1.52] | 1.73 [1.41 ; 1.88] | 4.0-10.0 | 0.04 | 0.04 | 0.26 | 0.09 | 0.05 | 0.04 | 0.26 |
| PMNs (Giga/l) | 0.14 [0.11 ; 0.28] | 0.16 [0.06 ; 0.23] | 0.2 [0.12 ; 0.25] | 0.37 [0.23 ; 0.58] | 1.5-7.0 | 0.07 | 0.07 | 0.25 | 0.21 | 0.05 | 0.08 | 0.50 |
| Monocyte (Giga/l) | 0.41 [0.28 ; 0.96] | 0.61 [0.33 ; 0.79] | 0.59 [0.52 ; 0.83] | 0.88 [0.66 ; 1.04] | 0.2-1.0 | 0.3 | 0.3 | 0.34 | 0.15 | 0.70 | 0.15 | 0.23 |
| Monocyte CD16- (%) | 61.16 [48.36 ; 85.34] | 36.87 [28.41 ; 78.05] | 63.48 [42.83 ; 70.75] | 31.61 [30.61 ; 44.08] | 80-85 | 0.25 | 0.25 | 0.62 | 0.15 | 0.42 | 0.70 | 0.21 |
| Monocyte Int ((%)/l) | 36.52 [13.12 ; 46.61] | 61.32 [17.61 ; 67.51] | 32.98 [24.74 ; 50.42] | 53.93 [53.03 ; 57.07] | 2-11 | 0.28 | 0.28 | 0.73 | 0.19 | 0.35 | 1.00 | 0.12 |
| Monocyte CD16+ (%) | 1.54 [0.91 ; 3.11] | 2.55 [1.58 ; 5] | 3.96 [2.3 ; 7.48] | 9.46 [1.48 ; 10.62] | 2-8 | 0.24 | 0.24 | 0.06 | 0.19 | 0.75 | 0.65 | 0.57 |
| HLA (AB/C) | 5094 [4173 ; 6396] | 4481.5 [3211.5 ; 8595] | 4208.5 [2956 ; 7247] | 7887 [4782 ; 8933] | 16884[5842-29175] | 0.47 | 0.47 | 0.86 | 0.13 | 0.75 | 0.39 | 0.16 |
| HLA CD16- (AB/C) | 3997.5 [2661 ; 6214] | 3071 [2552 ; 5782] | 3063.5 [2006.5 ; 5131] | 5029 [4800 ; 6781] |  | 0.62 | 0.62 | 0.57 | 0.44 | 0.81 | 0.25 | 0.38 |
| HLA Int (AB/C) | 7762.5 [4915.5 ; 10029] | 5624 [4515 ; 11591] | 7136.5 [4576 ; 12552] | 10730 [10487 ; 10881] |  | 0.68 | 0.68 | 1.00 | 0.19 | 0.70 | 0.44 | 0.44 |
| HLA CD16+ (AB/C) | 21660 [9066 ; 31767] | 16673 [10992 ; 29499] | 24749 [16673.5 ; 31627] | 47671 [39954 ; 53627] |  | 0.09 | 0.09 | 0.62 | 0.07 | 0.60 | 0.08 | 0.03 |
| Lymphocyte | 1.02 [0.53 ; 1.28] | 0.84 [0.55 ; 1.18] | 1.06 [0.91 ; 1.52] | 1.73 [1.41 ; 1.88] | 1.0-3.0 | 0.05 | 0.05 | 0.32 | 0.07 | 0.12 | 0.03 | 0.14 |
| LT4 (Giga/l) | 0.14 [0.11 ; 0.28] | 0.16 [0.06 ; 0.23] | 0.2 [0.12 ; 0.25] | 0.37 [0.23 ; 0.58] | 0.55-1.5 | 0.13 | 0.13 | 0.60 | 0.07 | 0.41 | 0.03 | 0.30 |
| LT4 (%) | 19.1 [10.4 ; 24.7] | 17.8 [8.7 ; 26.2] | 19.35 [12 ; 26.4] | 26 [14.6 ; 29.9] |  | 0.26 | 0.26 | 0.10 | 0.20 | 0.29 | 0.52 | 0.96 |
| LT8 (Giga/l) | 0.07 [0.03 ; 0.11] | 0.11 [0.05 ; 0.15] | 0.15 [0.11 ; 0.19] | 0.09 [0.06 ; 0.23] | 0.3-1.3 | 0.12 | 0.12 | 0.25 | 0.11 | 0.24 | 0.05 | 0.26 |
| LT8 (%) | 6.5 [5.3 ; 10.4] | 12.6 [10.8 ; 15.4] | 12.95 [8.15 ; 20.6] | 6.45 [3.3 ; 14] |  | 0.44 | 0.44 | 0.53 | 0.33 | 0.35 | 0.16 | 0.44 |
| LB (Giga/l) | 0.57 [0.31 ; 0.61] | 0.45 [0.28 ; 0.65] | 0.55 [0.35 ; 0.71] | 0.83 [0.5 ; 0.91] | 0.09-0.6 | 0.13 | 0.13 | 0.60 | 0.07 | 0.41 | 0.03 | 0.30 |
| LB (%) | 56.7 [50 ; 61.7] | 50.3 [45.9 ; 56.3] | 47.1 [41.45 ; 53.2] | 47.2 [40.8 ; 53.4] |  | 0.26 | 0.26 | 0.10 | 0.20 | 0.29 | 0.52 | 0.96 |
| L NK (Giga/l) | 0.1 [0.03 ; 0.16] | 0.11 [0.04 ; 0.19] | 0.15 [0.1 ; 0.3] | 0.31 [0.18 ; 0.43] | 0.15-1.1 | 0.12 | 0.12 | 0.25 | 0.11 | 0.24 | 0.05 | 0.26 |
| L NK (%) | 11.8 [8.8 ; 15.8] | 13.1 [7.1 ; 14.9] | 16.3 [9.8 ; 17.7] | 16.6 [11.3 ; 22] |  | 0.44 | 0.44 | 0.53 | 0.33 | 0.35 | 0.16 | 0.44 |
| CD4/CD8 | 4.05 [3.76 ; 5.7] | 3.81 [2.74 ; 5.6] | 2.73 [2.23 ; 4.92] | 2.92 [1.83 ; 4.31] |  | 0.42 | 0.42 | 0.38 | 0.33 | 0.20 | 0.23 | 0.89 |
| Treg (Giga/l) | 8.4 [6.1 ; 10.9] | 7.6 [6 ; 10.3] | 7.6 [7.1 ; 13.8] | 8.7 [6.5 ; 9] |  | 0.69 | 0.69 | 0.34 | 0.86 | 0.35 | 0.77 | 0.52 |
| PMN/Lymphocyte | 19.1 [10.4 ; 24.7] | 17.8 [8.7 ; 26.2] | 19.35 [12 ; 26.4] | 26 [14.6 ; 29.9] |  | 0.53 | 0.53 | 0.34 | 0.68 | 0.25 | 0.84 | 0.41 |

**Table S3.** Fluorescent-conjugated antibodies and reagents used for flow-cytometry.

|  |
| --- |
| BD Quantibrite™ Anti-HLA-DR/Anti-Monocyte kit<br>(anti-HLA-DR PE, clone L243 + anti-CD14 PerCP-Cy5.5, clone MφP9) |
| BD Quantibrite™ PE Fluorescence Quantitation Kit |
| BD Multitest™ 6-color TBNK Reagent with BD Trucount™ Tubes |
| CD16 APC-H7, clone 3G8 |
| CD3 - BB515 and APC, clone UCHT1 |
| IL-2 - PE, clone MQ1-17H12 |
| CD4 - PE-Cy7, clone SK3 |
| TNF-α - APC, clone MAb11 |
| IFN-γ - BV421, clone B27 |
| CD8 - PE-CF594, clone RPA-T8 |
| CD25- BV421, clone M-A251 |
| CD127 - PE, clone HIL-7R-M21 |
| Fixable Viability Stain 510 |

All from BD Biosciences, except CD3 – APC, from Biolegend.

